# Application of polygenic scores to a deeply phenotyped sample enriched for substance use disorders reveals extensive pleiotropy with psychiatric and medical traits

**DOI:** 10.1101/2024.01.22.24301615

**Authors:** Emily E. Hartwell, Zeal Jinwala, Jackson Milone, Sarah Ramirez, Joel Gelernter, Henry R. Kranzler, Rachel L. Kember

## Abstract

Co-occurring psychiatric, medical, and substance use disorders (SUDs) are common, but the complex pathways leading to such comorbidities are poorly understood. A greater understanding of genetic influences on this phenomenon could inform precision medicine efforts. We used the Yale-Penn dataset, a cross-sectional sample enriched for individuals with SUDs, to examine pleiotropic effects of genetic liability for psychiatric and medical traits. Participants completed an in-depth interview that provides information on demographics, environment, medical illnesses, and psychiatric and SUDs. Polygenic scores (PGS) for psychiatric disorders and medical traits were calculated in European-ancestry (EUR; n=5,691) participants and, when discovery datasets were available, for African-ancestry (AFR; n=4,918) participants. Phenome-wide association studies (PheWAS) were then conducted. In AFR participants, the only PGS with significant associations was bipolar disorder (BD), all of which were with substance use phenotypes. In EUR participants, PGS for major depressive disorder (MDD), generalized anxiety disorder (GAD), post-traumatic stress disorder (PTSD), schizophrenia (SCZ), body mass index (BMI), coronary artery disease (CAD), and type 2 diabetes (T2D) all showed significant associations, the majority of which were with phenotypes in the substance use categories. For instance, PGS_MDD_ was associated with over 200 phenotypes, 15 of which were depression-related (e.g., depression criterion count), 55 of which were other psychiatric phenotypes, and 126 of which were substance use phenotypes; and PGS_BMI_ was associated with 138 phenotypes, 105 of which were substance related. Genetic liability for psychiatric and medical traits is associated with numerous phenotypes across multiple categories, indicative of the broad genetic liability of these traits.

## Introduction

Medical illness and psychiatric disorders, including substance use disorders (SUDs), frequently co-occur. Individuals with chronic medical conditions are more likely to have a co-occurring SUD or psychiatric diagnosis[1–5] and over 9 million U.S. adults have a psychiatric disorder that co-occurs with an SUD[6]. The development of a comorbid disorder can exacerbate pre-existing conditions[7,8] and worsen an individual’s prognosis[9,10]. Moreover, co-occurring disorders can limit treatment options[11] and adversely affect treatment outcomes by reducing treatment adherence or decreasing its effectiveness[12–14]. Understanding the genetic underpinnings of comorbid disorders could improve their diagnosis, treatment, and ongoing management, thus informing precision medicine efforts.

Genetic liability for medical and psychiatric disorders has been discovered using genome-wide association studies (GWAS), which identify associations between common single nucleotide polymorphisms (SNPs) and the trait of interest. These studies have identified pleiotropic SNPs, i.e., those associated with multiple conditions. GWAS findings have also demonstrated significant genetic correlations between SUDs and other psychiatric disorders[15,16] and medical conditions[17]. These findings contribute to a growing body of evidence that shared genetic risk loci or common biological pathways may underlie co-occurring conditions.

Polygenic scores (PGS) provide a measure of an individual’s genetic risk for specific traits and as such are a complementary method to investigate genetic overlap. Previous studies have shown that PGS are a useful indicator of conditions such as cardiovascular disease[18], kidney disease[19], opioid use disorder[20], depression[21], and pain[22], among many others. PGS may also be used in phenome-wide association studies (PheWAS)[23] to provide insight into the pleiotropic nature of genetic liability for disorders[24,25]. PheWAS, which have been commonly implemented using electronic health record (EHR) databases, measure the association between a PGS for a disorder by testing it against multiple phenotypes in a hypothesis-free paradigm.

Here, we used the Yale-Penn sample—which comprises a diverse sample of participants recruited for genetic studies of cocaine, opioid, and alcohol dependence—to conduct PheWAS of psychiatric and medical PGS. Yale-Penn participants completed the Semi-Structured Assessment for Drug Dependence and Alcoholism (SSADDA) which queries medical, psychosocial, and substance use history and diagnoses, psychiatric diagnoses, and demographics[26,27]. Previous studies have utilized the Yale-Penn sample to conduct gene x environment studies[28], linkage and association studies of substance use and dependence[29–36], and to examine phenotypic associations[37]. These studies have shown shared genetic liability across SUDs, psychiatric disorders, and environmental traits.

Using the Yale-Penn sample, we created a simplified PheWAS dataset for genetic analysis and calculated PGS to examine pleiotropy for four major substance-related traits: alcohol use disorder, opioid use disorder, smoking initiation, and lifetime cannabis use[38]. PheWAS analyses in European-ancestry participants identified significant associations between SUD PGS and substance and psychiatric diagnoses and demographic and environmental phenotypes. Here, we extend this work by examining the associations of PGS for a variety of psychiatric and medical disorders in the Yale-Penn sample.

## Methods

### Participants and Procedures

The Yale-Penn sample (N=14,040) was recruited from five U.S. academic sites for studies of the genetics of cocaine, alcohol, and opioid use disorders. After they gave informed consent, all participants were administered the SSADDA and provided a blood or saliva sample for genotyping. The SSADDA comprises 24 modules that assess demographic information, environmental variables, medical history, and psychiatric and substance use history and diagnoses[26]. Additional information on data selection and cleaning has been published[38]. In brief, the SSADDA yields over 3,700 variables, which we refined to 691 variables for use in PheWAS. These variables are grouped into 25 categories: Demographics, Medical History, Substance Use (Tobacco, Alcohol, Cocaine, Opiate, Marijuana, Sedatives, Stimulants, Other drugs), Psychiatric (Major Depression, Conduct Disorder, Antisocial Personality Disorder [ASPD], Attention Deficit Hyperactivity Disorder [ADHD], Suicidality, Post-Traumatic Stress Disorder [PTSD], Generalized Anxiety Disorder [GAD], Panic Disorder, Social Phobia, Mania, Agoraphobia, Obsessive Compulsive Disorder [OCD], Schizophrenia, and Gambling) and Environment.

### Case and Control Definitions

Participants who endorsed Diagnostic and Statistical Manual (DSM) criteria for a given lifetime disorder (DSM-IV for psychiatric disorders, DSM-IV and DSM-5 for SUDs) were coded as cases and those who met no diagnostic criteria were considered controls. Participants meeting a sub-threshold number of criteria (e.g., one criterion when multiple are required for diagnosis) were excluded from analyses for that disorder. For individual symptoms (e.g., suicide attempt), participants who responded affirmatively were considered cases and those who did not were considered controls. When an item was not answered, participants were coded as “NA” and not included as either a case or a control for that variable.

### Genotyping and Imputation

In brief, Yale-Penn participants were genotyped in three batches using Illumina microarrays at Center for Inherited Disease Research (CIDR) or the Gelernter lab at Yale and imputed using the Michigan Imputation Server[39] with the 1000 Genomes phase 3 reference panel[40]. Details on genotyping, imputation, and quality control for the genetic data have previously been reported[36,38,41,42].

Ancestry-specific PGS were calculated using PRS-Continuous Shrinkage (PRS-CS) software[43] from GWAS in discovery samples for anorexia (AN)[44], autism spectrum disorder (ASD)[45], bipolar disorder (BD)[46], generalized anxiety disorder (GAD)[47], major depressive disorder (MDD)[48], obsessive compulsive disorder (OCD)[49], panic disorder (PD)[50], post-traumatic stress disorder (PTSD)[51], schizophrenia (SCZ)[52], Tourette syndrome (TS)[53], body mass index (BMI)[54], coronary artery disease (CAD)[55] and type 2 diabetes (T2D)[56] (Supplementary Table 1). All GWAS were available for European-ancestry (EUR), but only BD[57], GAD[47], MDD[21], PTSD[51], and SCZ[52] were available for African-ancestry (AFR). Discovery GWAS were selected based on their public availability and excluded the Yale-Penn sample.

### Statistical Analysis

For PGS with available primary phenotypes (diagnoses for AN, ASD and TS are not available in the Yale-Penn sample), we tested for association between the PGS and the primary phenotype. We next conducted a series of PheWAS using logistic regression models for binary traits and linear regression models for continuous traits, adjusting for age, sex, and the top 10 principal genetic components within each ancestry. Phenotypes in which there were less than 100 cases or controls were excluded. For available phenotypes, a second PheWAS was run that covaried for the primary diagnostic phenotype. A Bonferroni correction was applied to each ancestry group to account for multiple comparisons (AFR phenotypes n=574, p=8.7×10^−05^; EUR phenotypes n=620, p=8.1×10^−05^).

## Results

### Sample

Genetic data were available for 10,275 of the 14,040 participants, the majority of whom (54.46%) were male. The sample included 4,851 AFR participants (55.2% males) and 5,424 EUR participants (51.1% males) whose mean ages were 41.47 (SD=10.16) and 39.79 (SD=12.91), respectively. Supplementary Table 2 shows demographic information by for available primary phenotypes.

### Primary phenotypic associations of PGS

For the PGS with primary phenotypes available, we tested the association of PGS with each primary phenotype (Figure 1A). In AFR participants, none of the PGS were associated with their primary phenotype. In EUR participants, PGS for three psychiatric disorders (PGS_MDD_, PGS_PD_, and PGS_PTSD_) and three medical traits (PGS_BMI_, PGS_CAD_ and PGS_T2D_) were associated with their primary phenotype at a *p*-value of <0.05.

**Figure 1:**
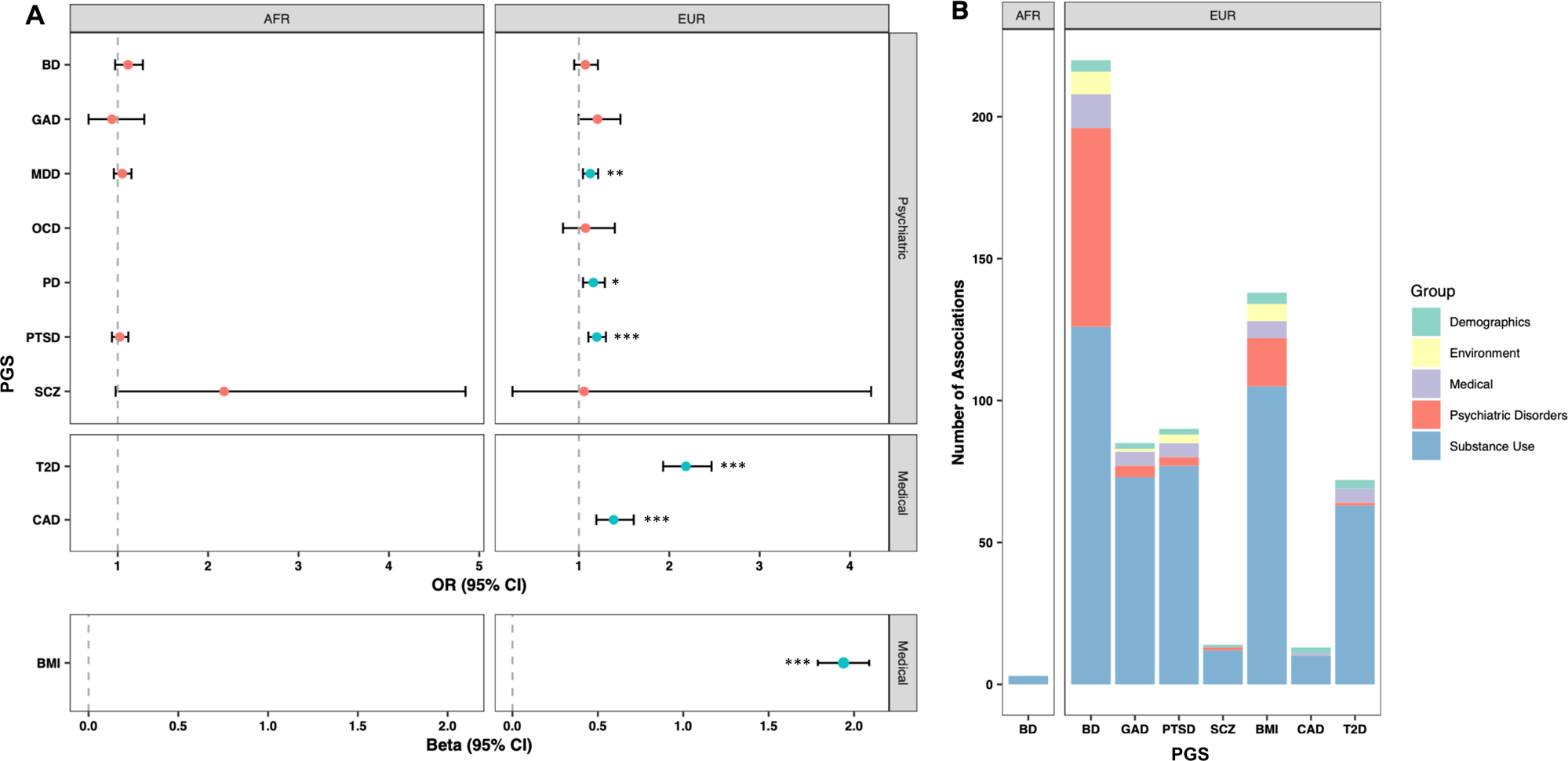
Primary and secondary associations of psychiatric and medical PGS. A: Effect size and 95% confidence intervals for associations between PGS and their corresponding primary phenotype, if available. Asterisks indicate p-value for significant associations: *p<0.05, ** p<0.01, *** p<0.001. B: Number of associations within each category for the PGS with significant associations. BD: Bipolar Disorder, GAD: generalized anxiety disorder, MDD: major depressive disorder, OCD: obsessive compulsive disorder, PD: panic disorder, PTSD, post-traumatic stress disorder, SCZ: schizophrenia, T2D: diabetes, CAD: coronary artery disease, BMI: body mass index.

We next examined phenotypic associations of each PGS other than the primary phenotype. In AFR participants, after Bonferroni correction, there were significant associations for PGS_BD_ (Supplementary Table 3). No other associations were observed among AFR participants following Bonferroni correction (Supplementary Tables 4-7). In EUR participants, there were significant associations for five of the psychiatric disorders (PGS_MDD_, PGS_GAD_, PGS_PTSD_, PGS_SCZ_, and PGS_TS_) and medical traits (PGS_BMI_, PGS_CAD_ and PGS_T2DM_), whereas there were no significant associations for PGS_BD_, PGS_AN_, PGS_ASD_, PGS_OCD_ or PGS_PD_ (Supplementary Tables 8-20).

### Phenome-wide analysis of psychiatric PGS

#### Bipolar Disorder (BD)

In AFR participants, PGS_BD_ was associated with three phenotypes in the substance use category, all related to cocaine (e.g., regularly use cocaine; OR=1.14, *p*=8.6×10^−5^; Supplementary Tables 3 and 21). PGS_BD_ was not associated with any phenotypes in EUR participants (Supplementary Table 8).

#### Major Depressive Disorder (MDD)

In EUR participants, PGS_MDD_ was associated with 220 phenotypes across 17 categories (Figures 1B and 2; Supplementary Tables 9 and 21). Although PGS_MDD_ was not significantly associated with the MDD diagnosis following Bonferroni correction (OR=1.13, *p*=3.8×10^−3^; Figure 1A), it was significantly associated with 15 phenotypes in the depression category, most significantly the MDD criterion count (β=0.40, *p*=3.0×10^−15^). These phenotypes remained significantly associated when covarying for MDD diagnosis (Supplementary Figure 1, Supplementary Table 9).

**Figure 2:**
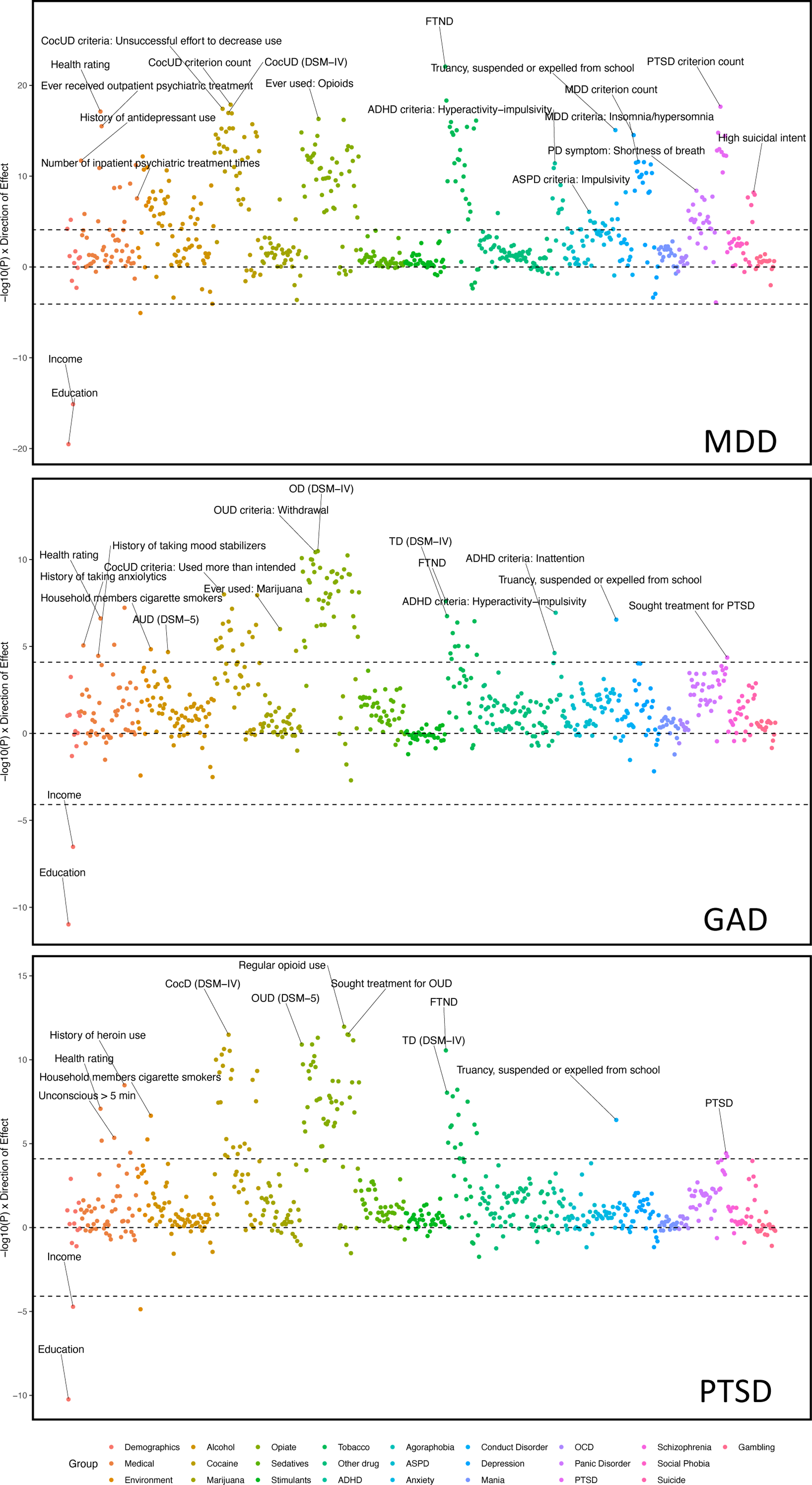
Phenome-wide association results for MDD, GAD and PTSD. Phenotype categories are plotted along the x-axis, and –1log10 p-value x direction of effect is plotted on the y-axis. Selected phenotypes passing Bonferroni correction are labelled.

PGS_MDD_ also showed 55 significant associations with other psychiatric disorders. Notably, PGS_MDD_ was the only PGS associated with any phenotypes in the depression, generalized anxiety, panic disorder, agoraphobia, and suicide categories (Supplementary Table 21). In the anxiety disorder categories, the panic disorder symptom shortness of breath (OR=1.29, *p*=4.0×10^−9^), PTSD criterion count (β=0.26, *p*=2.3×10^−18^), and the sum of physical reactions (β=0.14, *p*=5.9×10^−6^) for GAD were the most significant. Five phenotypes related to suicide were significantly associated with PGS_MDD_, including high suicidal intent (OR=1.37, *p*=6.5×10^−9^) and suicide attempt (OR=1.29, *p*=2.2×10^−8^). Covarying for the MDD diagnosis reduced the number of associations in the panic disorder category from 18 to 13, and the number of associations with GAD from 4 to 0. Other categories with significant associations included those with conduct disorder (e.g., truancy, being suspended or expelled from school (OR=1.31, *p*=8.5×10^−16^)), ASPD (e.g., impulsivity (OR=1.17, *p*=8.6×10^−7^)), and ADHD (e.g., criterion count (β=0.09, *p*=9.6×10^−10^)). Additionally, PGS_MDD_ was significantly associated with demographic and environmental phenotypes, including negatively with education (β=−0.10, *p*=8.7×10^−20^) and household income (β=−0.26, *p*=7.6×10^−16^), and positively with childhood adversity (OR=1.29, *p*=6.8×10^−13^). PGS_MDD_ was the only PGS associated with the number of inpatient psychiatric treatments (β=0.50, *p*=2.8×10^−8^), emotional problems (OR=1.28, *p*=6.0×10^−12^), and history of antidepressant use (OR=1.24, *p*=2.0×10^−12^).

PGS_MDD_ was also significantly associated with 126 substance use phenotypes, 122 of which remained significant after covarying for the MDD diagnosis. The substance use traits most significantly associated with PGS_MDD_ in each category were the Fagerström Test for Nicotine Dependence (FTND) score (β=0.38, *p*=8.7×10^−23^), criterion count for DSM-5 cocaine use disorder (CocUD; β=0.56, *p*=1.3×10^−18^), “ever used” opioids (OR=1.31, *p*=5.1×10^−17^), and DSM-IV alcohol abuse (OR=1.25, *p*=2.3×10^−11^). Notably, PGS_MDD_ had the most alcohol associations of the PGS tested.

#### Generalized Anxiety Disorder (GAD)

PGS_GAD_ was associated with 85 phenotypes in EUR participants (Figures 1B and 2; Supplementary Tables 10 and 21). Although it was not significantly associated with the primary diagnosis of GAD (OR=1.21, *p*=0.06; Figure 1), it was the only PGS to be associated with a history of anxiolytic treatment (OR=1.19, *p*=8.7×10^−6^).

PGS_GAD_ was significantly associated with four phenotypes related to other psychiatric disorders, which included hyperactivity-impulsivity (β=0.18, *p*=1.2×10^−7^) and inattention (β=0.17, p=2.4×10^−5^) for ADHD; truancy, being suspended or expelled from school (OR=1.18, *p*=2.9×10^−7^) for conduct disorder; and seeking treatment for PTSD (OR=1.21, *p*=4.4×10^−5^). Additionally, PGS_GAD_ was significantly associated with non-psychiatric phenotypes, such as health rating (higher value indicates poorer health; β=0.07, *p*=2.4×10^−7^) and household members being cigarette smokers (OR=1.15, *p*=1.5×10^−5^), and negatively with education (β=−0.07, *p*=1.0×10^−11^) and household income (β=−0.16, *p*=3.0×10^−7^).

PGS_GAD_ was significantly associated with 73 substance use phenotypes, particularly in the tobacco, cocaine, and opioid categories (e.g., DSM-IV opioid dependence (OR=1.23, *p*=3.3×10^−11^), using more cocaine than intended (OR=1.18, *p*=1.0×10^−8^), FTND score (β=0.21, *p*=2.5×10^−8^), and DSM-5 alcohol use disorder (AUD, OR=1.16, *p*=2.1×10^−5^)). Unlike other psychiatric PGS, PGS_GAD_ also had two significant associations with marijuana use.

Covarying for the primary phenotype, half of the phenotypes associated with PGS_GAD_ were not significant, although the association with anxiolytic treatment remained significant (Supplementary Figure 2, Supplementary Table 10). The tobacco category had the greatest reduction in number of associations, with 8 of 11 phenotypes no longer significant when GAD diagnosis was included as a covariate.

#### Post-traumatic stress disorder (PTSD)

PGS_PTSD_ was associated with a total of 90 phenotypes in EUR participants (Figures 1B and 2; Supplementary Tables 11 and 21). PGS_PTSD_ showed significant associations with the diagnosis of PTSD (OR=1.20, *p*=3.8×10^−5^) and with treatment seeking for PTSD (OR=1.21, *p*=5.8×10^−5^). The only other association in the psychiatric category was with truancy, being suspended, or expelled from school in the conduct disorder category (OR=1.18, *p*=3.9×10^−7^). These associations were no longer significant when the PTSD diagnosis was used as a covariate in the analysis (Supplementary Figure 3, Supplementary Table 11).

PGS_PTSD_ was significantly associated with 77 cocaine, tobacco, and opioid use phenotypes, including DSM-IV dependence and withdrawal symptoms for all three substances. Almost half of these traits were not significant when the PTSD diagnosis was used as a covariate.

PGS_PTSD_ was also negatively associated with education (β=−0.07, *p*=5.8×10^−11^) and household income (β=−0.13, *p*=1.9×10^−5^). In the environment category, PGS_PTSD_ was associated with household members being cigarette smokers (OR=1.18, *p*=2.2×10^−7^), exposure to violent crime before the age of 13 (OR=1.23, *p*=5.6×10^−6^), and negatively with the parent being the primary caregiver (OR=0.82, *p*=1.4×10^−5^). However, none of the environmental phenotypes remained significant when the PTSD diagnosis was used as a covariate. PGS_PTSD_ was also associated with health rating (β=0.08, *p*=8.4×10^−8^) and seeking outpatient psychiatric treatment (OR=1.15, *p*=6.7×10^−6^).

#### Schizophrenia (SCZ)

PGS_SCZ_ was associated with 14 phenotypes in EUR participants (Supplementary Figure 4; Supplementary Tables 12 and 21). The only associations with non-substance use phenotypes were with truancy, being suspended or expelled from school (OR=1.21, *p*=3.4×10^−7^) in the conduct disorder category and negatively with household income (β=−0.15, *p*=4.4×10^−5^) in the demographics section.

Among substance use phenotypes, PGS_SCZ_ was significantly associated with several alcohol use phenotypes (e.g., reduction in other activities (OR=1.22, *p*=4.2×10^−9^) and failure to fulfill obligations (OR=1.20, *p*=1.0×10^−7^)). PGS_SCZ_ was also associated with cocaine use phenotypes, such as failure to fulfill obligations (OR=1.16, *p*=1.4×10^−5^).

#### Tourette’s Syndrome (TS)

In EUR participants, PGS_TS_ was associated with 1 environmental phenotype (Supplementary Tables 13 and 21), frequency of moving/relocation as a child (β=0.18, *p*=7.8×10^−5^).

### Phenome-wide analysis of medical PGS

#### Body-mass index (BMI)

In EUR participants, PGS_BMI_ was associated with 138 phenotypes (Figures 1B and 3; Supplementary Table 18 and 21), which was most significant for the primary phenotype, BMI (β=1.94, *p*=4.1×10^−133^). Education (β=−0.11, *p*=1.2×10^−21^), household income (β=−0.19, *p*=1.6×10^−8^), and number of children (β=0.11, *p*=1.2×10^−8^) were other demographic variables with significant associations. PGS_BMI_ was associated with 6 medical phenotypes, including health rating (β=0.13, *p*=5.9×10^−17^), diabetes (OR=1.67, *p*=2.6×10^−12^), and number of medical problems (β=0.15, *p*=9.6×10^−11^). The demographic phenotypes remained associated when BMI was used as a covariate, but associations with health rating, diabetes and medical problems did not (Supplementary Figure 5, Supplementary Table 18).

**Figure 3:**
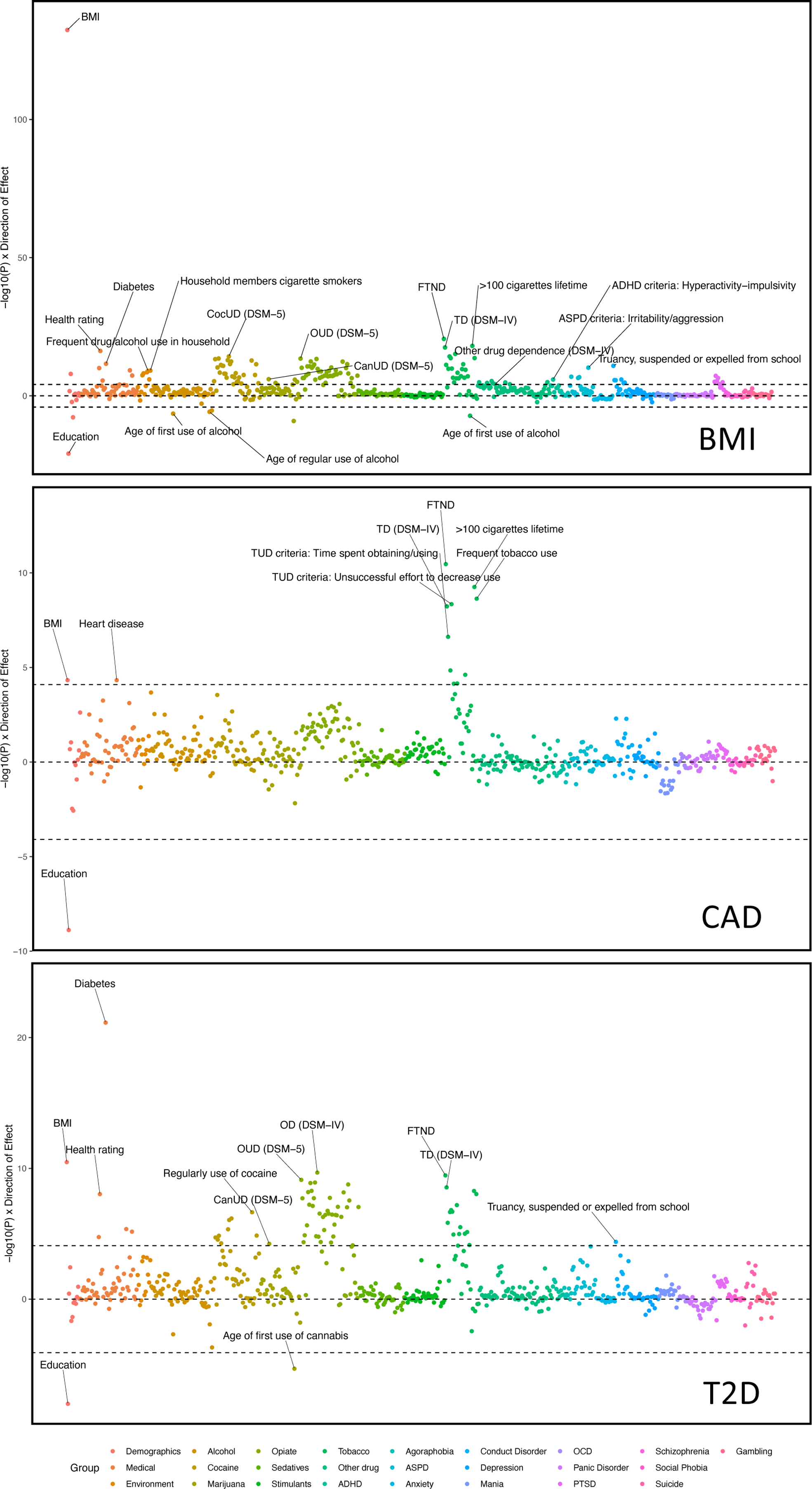
Phenome-wide association results for BMI, CAD and T2D. Phenotype categories are plotted along the x-axis, and –1log10 p-value x direction of effect is plotted on the y-axis. Selected phenotypes passing Bonferroni correction are labelled.

Seventeen psychiatric disorder phenotypes were associated with PGS_BMI_, including 10 associations with traits in the conduct disorder and ASPD categories including truancy, being suspended or expelled from school (OR=1.27, *p*=1.5×10^−11^) and irritability/aggression (OR=1.24, *p*=6.8×10^−11^), respectively; hyperactivity-impulsivity (β=0.19, *p*=1.3×10^−6^) in the ADHD section; and six phenotypes in the PTSD section, including reexperiencing (OR=1.21, *p*=6.4×10^−8^) and PTSD diagnosis (OR=1.24, *p*=1.3×10^−5^). When covarying for BMI, only 5 of the 17 associations remained significant.

PGS_BMI_ was also significantly associated with 105 substance use phenotypes, including the FTND score (β=0.40, *p*=2.6×10^−21^), DSM-5 CocUD (OR=1.30, *p*=5.7×10^−15^), DSM-5 opioid use disorder (OUD, OR=1.29, *p*=3.5×10^−14^), and DSM-5 cannabis use disorder (CanUD, OR=1.19, *p*=9.9×10^−7^). There were also numerous associations of PGS_BMI_ with heaviness of use, withdrawal, and physiological symptoms for a variety of substances. Although PGS_BMI_ was not associated with an AUD diagnosis, it was uniquely negatively associated with the ages of first alcohol use (β=−0.27, *p*=3.4×10^−7^), regular use (β=−0.28, *p*=1.6×10^−6^), and first intoxication (β=−0.25, *p*=4.6×10^−6^). The alcohol phenotypes were no longer significant when BMI was included as a covariate, although the majority of the other substance use phenotypes remained significantly associated.

PGS_BMI_ was also associated with several environmental variables. These included exposures to substance use in childhood, such as household members being cigarette smokers (OR=1.25, *p*=7.8×10^−10^), frequent drug/alcohol use in the household (OR=1.23, *p*=1.0×10^−9^), and awareness of household members using substances (OR=1.17, *p*=1.2×10^−6^). Lifetime trauma assessment (OR=1.20, *p*=7.9×10^−9^) and childhood adversity (OR=1.23, *p*=3.6×10^−8^) were also significantly associated with PGS_BMI_.

#### Coronary artery disease (CAD)

In EUR participants, PGS_CAD_ was significantly associated with 13 phenotypes (Figures 1B and 3; Supplementary Tables 19 and 21), including the primary phenotype of heart disease (OR=1.38, *p*=4.7×10^−5^), BMI (β=0.30, *p*=4.7×10^−5^) and a negative association with education (β=−0.07, *p*=1.3×10^−9^). The remaining significant associations were with 10 tobacco use phenotypes, including FTND score (β=0.25, *p*=3.4×10^−11^) and unsuccessful efforts to reduce smoking (OR=1.19, *p*=4.5×10^−9^). The majority of these associations remained significant when the primary phenotype was included as a covariate (Supplementary Figure 6; Supplementary Table 19).

#### Type 2 Diabetes (T2D)

PGS_T2D_ in EUR participants was significantly associated with 71 phenotypes, including diabetes (OR=2.18, *p*=7.2×10^−22^) (Figure 1B and 3; Supplementary Tables 20 and 21). PGS_T2D_ was associated with seven medical and demographic phenotypes, including BMI (β=0.58, *p*=3.3×10^−11^), health rating (β=0.10, *p*=9.3×10^−9^) and number of medical problems (β=0.11, *p*=1.8×10^−5^), and negatively with education (β=−0.07, *p*=9.9×10^−9^). Only the number of medical problems was not significant when the primary phenotype was included as a covariate.

Truancy, being suspended or expelled from school in the conduct disorder group was the only psychiatric phenotype associated with PGS_T2D_ (OR=1.17, *p*=4.2×10^−5^). PGS_T2D_ was significantly associated with 63 substance use phenotypes, including the FTND score (β=0.29, *p*=3.5×10^−10^), DSM-5 OUD (OR=1.25, *p*=7.6×10^−10^), DSM-5 CocUD (OR=1.20, *p*=8.9×10^−7^), and DSM-5 CanUD (OR=1.17, *p*=5.9×10^−5^). The majority of these remained significant when covarying for the primary phenotype (Supplementary Figure 7, Supplementary Table 20).

## Discussion

This study examined the performance of psychiatric and medical PGS in the deeply-phenotyped Yale-Penn sample, in which most participants were ascertained based on having one or more lifetime SUDs. The SSADDA yields a wealth of phenotypic data not typically available in EHR-based biobanks traditionally used for this type of analysis, therefore we were able to both replicate previous findings and identify several novel cross-trait associations. For all PGS, the largest number of associations were with phenotypes in the substance use categories. This is consistent with the high prevalence of SUDs and the large number of individual traits ascertained for each substance in this sample, and highlights the high degree of pleiotropy of SUDs with both psychiatric and medical phenotypes. Also, as might be expected, compared to the medical PGS, psychiatric disorder PGS showed more associations with phenotypes in psychiatric categories, both within and cross disorder.

Several PGS were significantly associated with their primary phenotypes. The three medical PGS were associated with their respective primary phenotypes, indicative of the power of the PGS. PGS_PTSD_ in EUR participants was associated with a PTSD diagnosis. However, PGS for MDD and GAD in EUR participants were not associated with their respective primary diagnosis following Bonferroni correction, though both were associated with related phenotypes, such as DSM criterion count for MDD and the use of medications to treat anxiety. The lack of association of PGS with their primary phenotypes could be due to the sample’s ascertainment strategy, which focused on the presence of one or more SUDs.

Some PGS did not yield any significant associations. In AFR participants, only the PGS_BD_ showed any significant associations and none were BD-related phenotypes. Although no other associations with PGS were significant, some of the AFR PGS showed nominal associations (i.e., p<0.05) that may become significant with a better powered PGS derived from a larger originating GWAS (e.g., the association of PGS_MDD_ with “ever depressed”). Because the SSADDA interview does not assess autism, Tourette’s Syndrome, or eating disorders, primary associations for these PGS could not be tested.

PGS_MDD_ showed the most associations of any PGS tested. Notably, it was also the only PGS to yield significant associations with depression- and suicide-related phenotypes. While other EHR-based PheWAS have demonstrated strong associations of MDD with the primary diagnosis [58,59], our strongest association among depression phenotypes was for the MDD criterion count. Moreover, each of the individual nine MDD diagnostic criteria were also significantly associated with PGS_MDD_, suggesting a genetic contribution to each. Most of the associations with psychiatric phenotypes remained significant when the depression diagnosis was covaried, indicating that the associations are not due to co-occurring MDD. As with previous findings in an EHR-based PheWAS, we observed associations of PGS_MDD_ with alcohol and tobacco use phenotypes, GAD, PTSD, and agoraphobia[58]. The association between SUDs and MDD was also found in our previous analysis in this sample, which demonstrated associations between PGS for SUDs and a number of depression phenotypes[38]. Interestingly, numerous withdrawal-related phenotypes for cocaine, tobacco, opioids, and alcohol were also significantly associated with PGS_MDD_, as were treatment-seeking for depression and other psychiatric disorders.

Few studies have examined the performance of anxiety-related PGS. One study in which a PGS_PTSD_ was tested in four EHR-based biobanks[60] showed significant associations with a PTSD diagnosis, a SUD diagnosis, and tobacco dependence, as well as numerous associations with medical conditions, including circulatory and respiratory diseases. In contrast to our findings, that study showed associations with various anxiety disorders and depression, which may have been due to the large size of the included biobanks and the higher number of cases for anxiety disorders. Our PheWAS results for both PGS_PTSD_ and PGS_GAD_ were associated with cocaine, opioid, and tobacco diagnoses, criterion counts, and treatment seeking for use of those substances, which is suggestive of an association of greater genetic risk for anxiety with greater SUD severity.

Participants who, during screening for study participation, self-reported having a schizophrenia or bipolar disorder diagnosis were excluded from the Yale-Penn sample. Thus, the lack of associations for PGS_SCZ_ and PGS_BD_ with the primary diagnoses and related phenotypes was not unexpected. As with previous PheWAS, PGS_SCZ_ was associated with substance use and personality disorder phenotypes[61]. Given the high rate at which SCZ and tobacco use co-occur, and previously observed association of PGS for SCZ with tobacco use[61], the lack of associations here were unexpected and may also be attributable to the exclusion of participants with psychotic disorders from the Yale-Penn sample.

In addition to all three medical PGS being strongly associated with their primary phenotype, they were associated with BMI and several tobacco-related phenotypes. Previous studies conducted using data from the UK Biobank and Penn Medicine BioBank showed associations of PGS_BMI_ with T2D, circulatory system disorders, and sleep problems[25,62]. We also found associations of the PGS_BMI_ with numerous substance-related phenotypes and environmental factors. Lifetime trauma assessment, childhood adversity, and childhood exposure to substance use were also significantly associated with PGS_BMI,_ experiences that have been shown to predict higher BMI[63]. PGS_T2D_, as expected, was associated with measures of poor health and numerous substance use phenotypes, the majority of which persisted after controlling for a diabetes diagnosis. Higher rates of SUDs have been observed in individuals with T2D[64] and individuals with a SUD and T2D experience poorer medical outcomes and higher mortality than those with T2D alone[65], though little is currently known about pleiotropy of these traits. Akin to previous work[66], PGS_CAD_ was associated with tobacco use phenotypes but no other substance use, such as alcohol phenotypes, or medical disorders, such as T2D. PGS_CAD_ did not yield any associations in the psychiatric category and PGS_T2D_ only had one, which was no longer significant covarying for diabetes diagnosis, suggesting that genetic liability for these medical disorders is not associated with psychiatric phenotypes in this sample.

This study should be interpreted in light of the strengths and limitations. The Yale-Penn dataset used as a target sample is comparatively small and cross-sectional, without longitudinal data and medical records data available in large, EHR-based genetic studies. However, the in-depth SSADDA interview provides granular psychiatric and substance use data not available in EHR-based biobanks, which provide the possibility of novel insights into the pleiotropy of co-occurring traits. The Yale-Penn sample excluded individuals with certain severe mental illnesses, thus limiting our ability to observe some associations, e.g., PGS_SCZ_ was not associated with the primary diagnosis of schizophrenia. Available discovery GWAS varied in size and those that included individuals of AFR ancestry GWAS were not available for all the phenotypes of interest. Because the Yale-Penn sample includes similar numbers of AFR and EUR participants, we believe that larger discovery GWAS in AFR participants and the accompanying increase in statistical power will be more informative of pleiotropy in non-EUR populations.

Despite these limitations, our findings demonstrate the pleiotropic nature of genetic liability for psychiatric and medical disorders. Both psychiatric and medical PGS were broadly associated with substance use phenotypes in a sample enriched for individuals with SUDs. Despite the extensive pleiotropy found, we also identified associations that were unique to specific PGS. Furthermore, psychiatric PGS were more likely to be associated with psychiatric disorders compared to medical PGS, suggesting some level of specificity of genetic architecture within categories. Many phenotypes remained associated when covarying for the primary phenotype on which the PGS was based, suggesting that the genetic liability for the disorders in question is the primary driver of the associations. Overall, we find evidence that genetic liability for psychiatric and medical disorders partially underlies the common co-occurrence of these traits with SUDs.

## Data Availability

All data produced in the present study are available upon reasonable request to the authors

## Author contributions

E.E.H and R.L.K conceived and designed the study; Z.J, J.M, S.R analyzed the data; J.G and H.R.K acquired the data; E.E.H, Z.J, J.M, S.R, R.L.K interpreted the data and drafted the manuscript; all authors revised the manuscript for intellectual content and provided final approval for submission.

## Funding

This study was funded by Department of Veterans Affairs grants IK2 CX002336 (EEH), I01 BX004820 (HRK), VISN1 MIRECC (JG), and the VISN4 MIRECC (HRK, RLK, EEH), and NIH grants R01DA037974 (JG), R01DA058862 (JG), K01AA028292 (RLK)

## Competing interests

Dr. Kranzler is a member of advisory boards for Dicerna Pharmaceuticals, Sophrosyne Pharmaceuticals, Enthion Pharmaceuticals, and Clearmind Medicine; a consultant to Sobrera Pharmaceuticals; the recipient of research funding and medication supplies for an investigator-initiated study from Alkermes; and a member of the American Society of Clinical Psychopharmacology’s Alcohol Clinical Trials Initiative, which was supported in the last three years by Alkermes, Dicerna, Ethypharm, Lundbeck, Mitsubishi, Otsuka, and Pear Therapeutics. Drs. Gelernter and Kranzler hold U.S. patent 10,900,082 titled: “Genotype-guided dosing of opioid agonists,” issued 26 January 2021.

## Notes

### Author Declarations

University of Pennsylvania Institutional Review Board gave ethical approval for this work.

