## Supplementary Figures for "Application of polygenic scores to a deeply phenotyped sample enriched for substance use disorders reveals extensive pleiotropy with psychiatric and medical traits"

Supplementary Figure 1: $PGS_{MDD}$ covarying for MDD

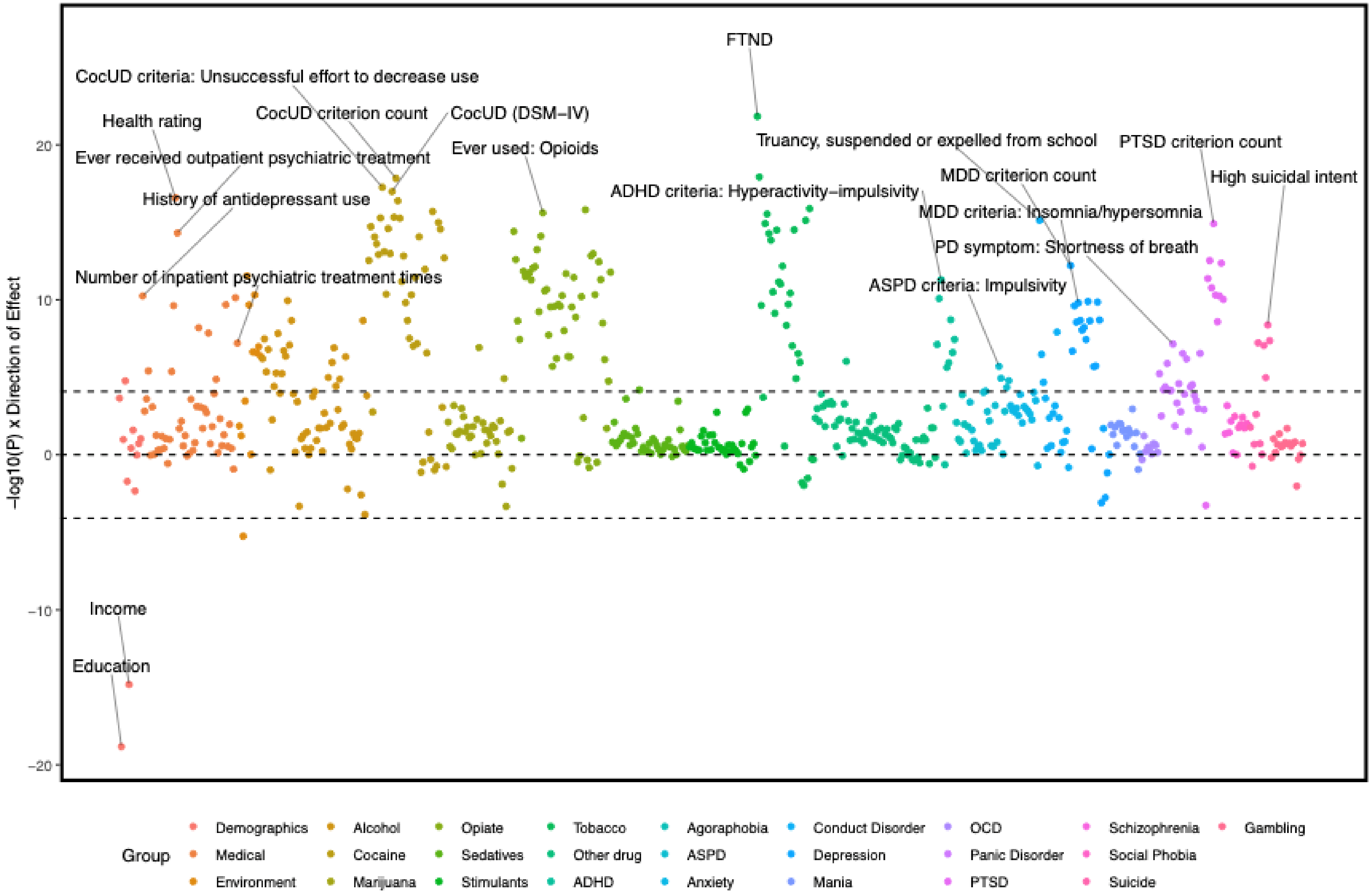

Supplementary Figure 2: $PGS_{GAD}$ covarying for GAD

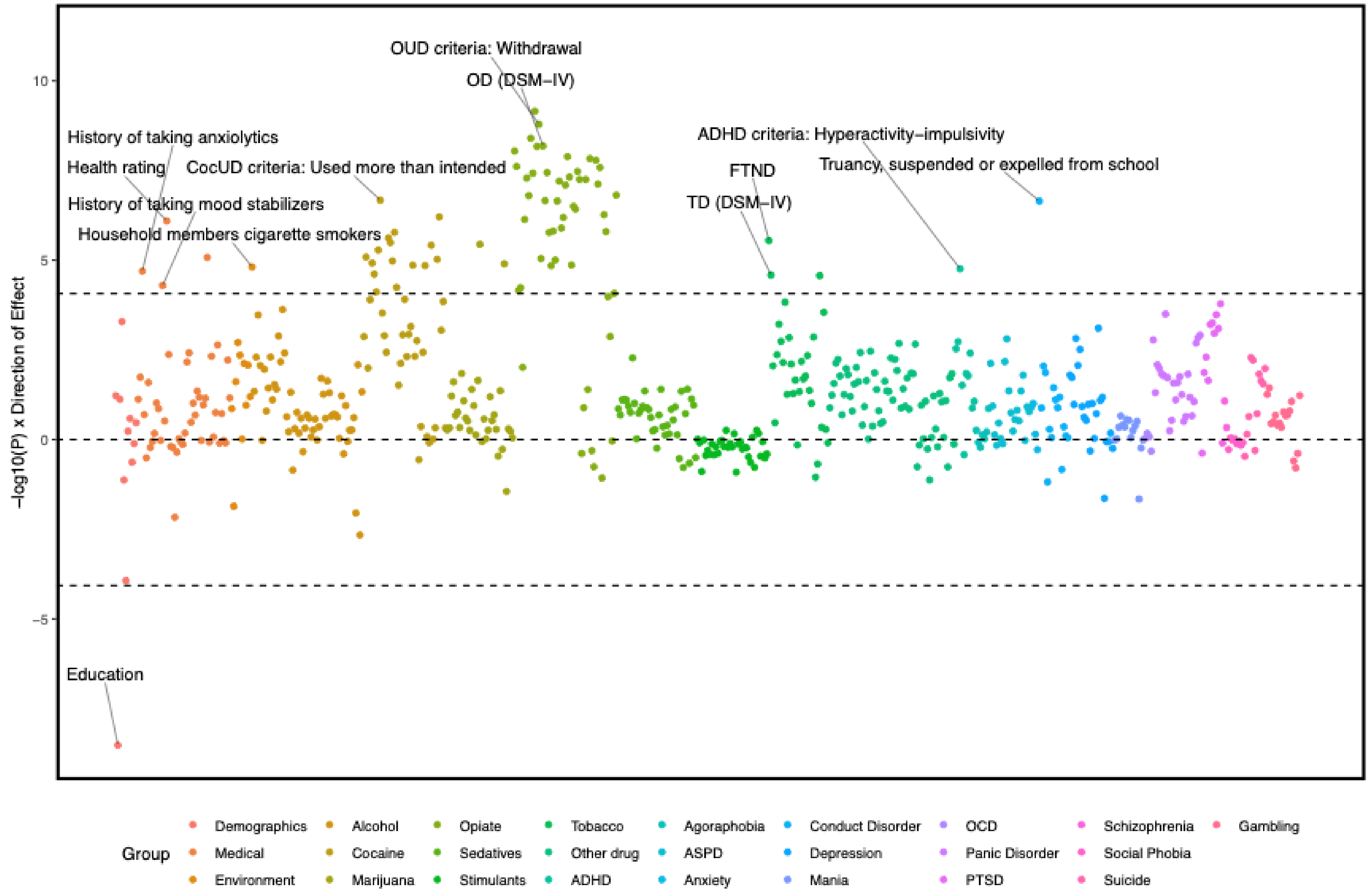

Supplementary Figure 3: $PGS_{PTSD}$ covarying for PTSD

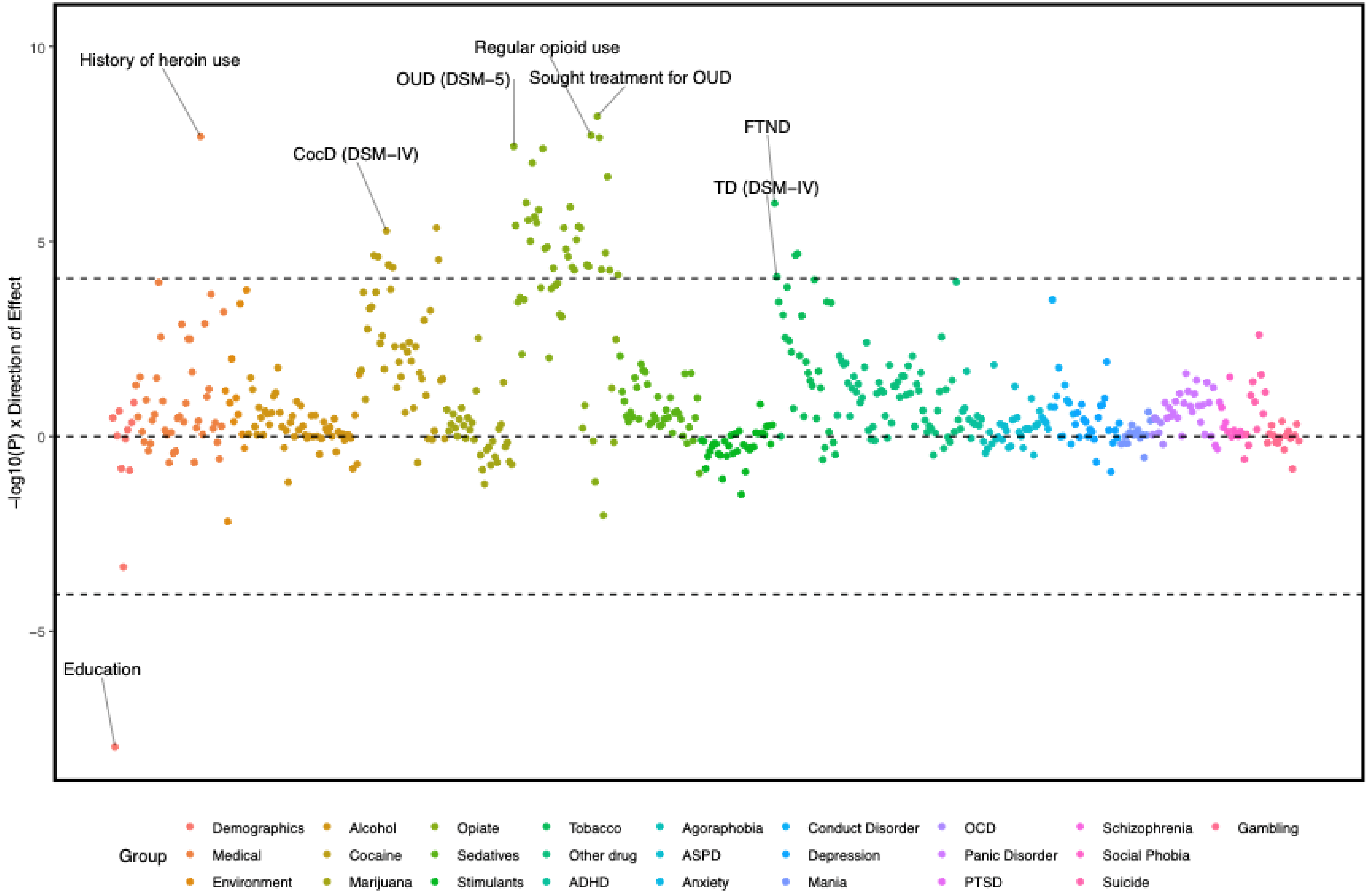

Supplementary Figure 4: $PGS_{SCZ}$

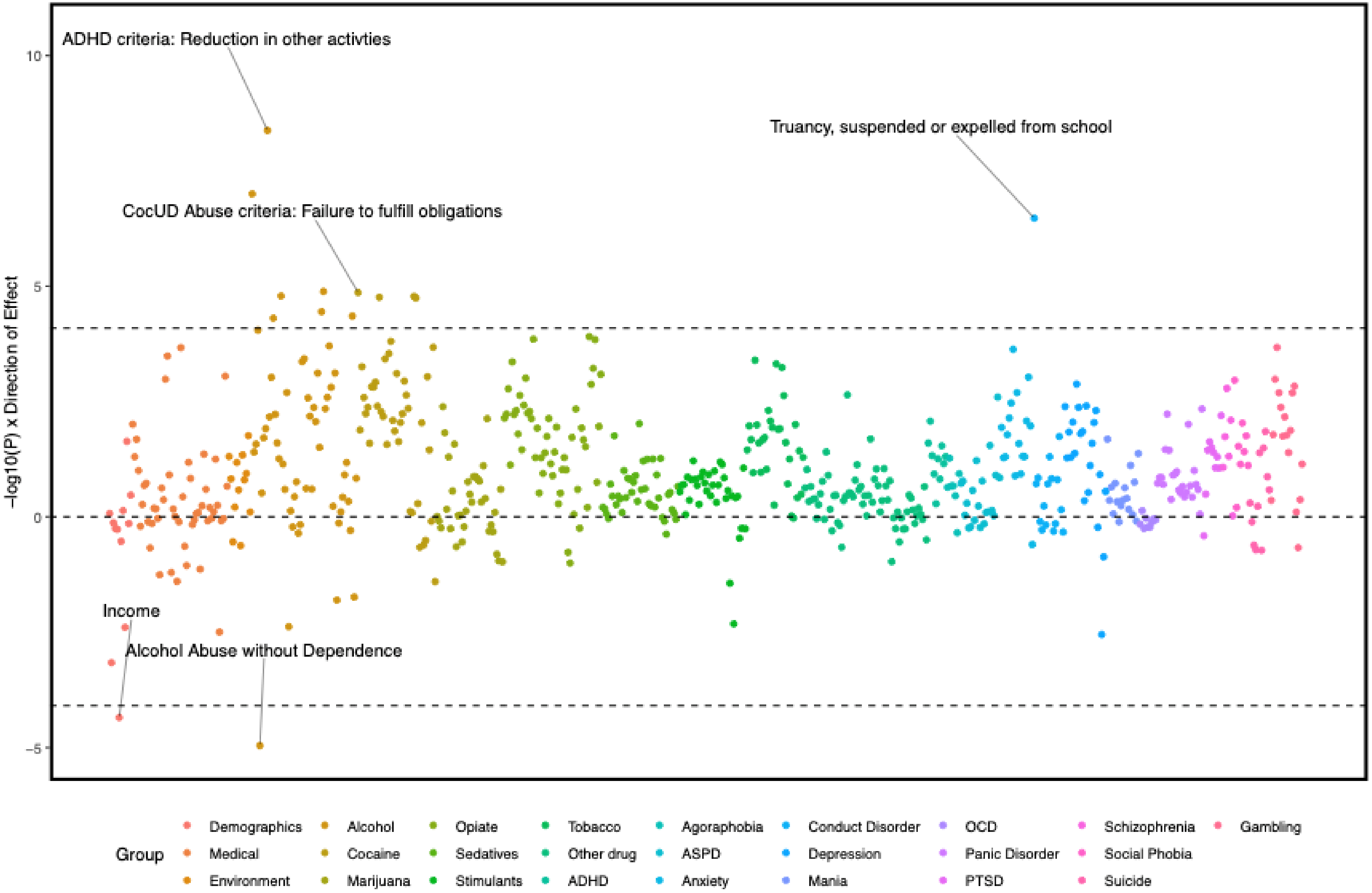

Supplementary Figure 5: $PGS_{BMI}$ covarying for BMI

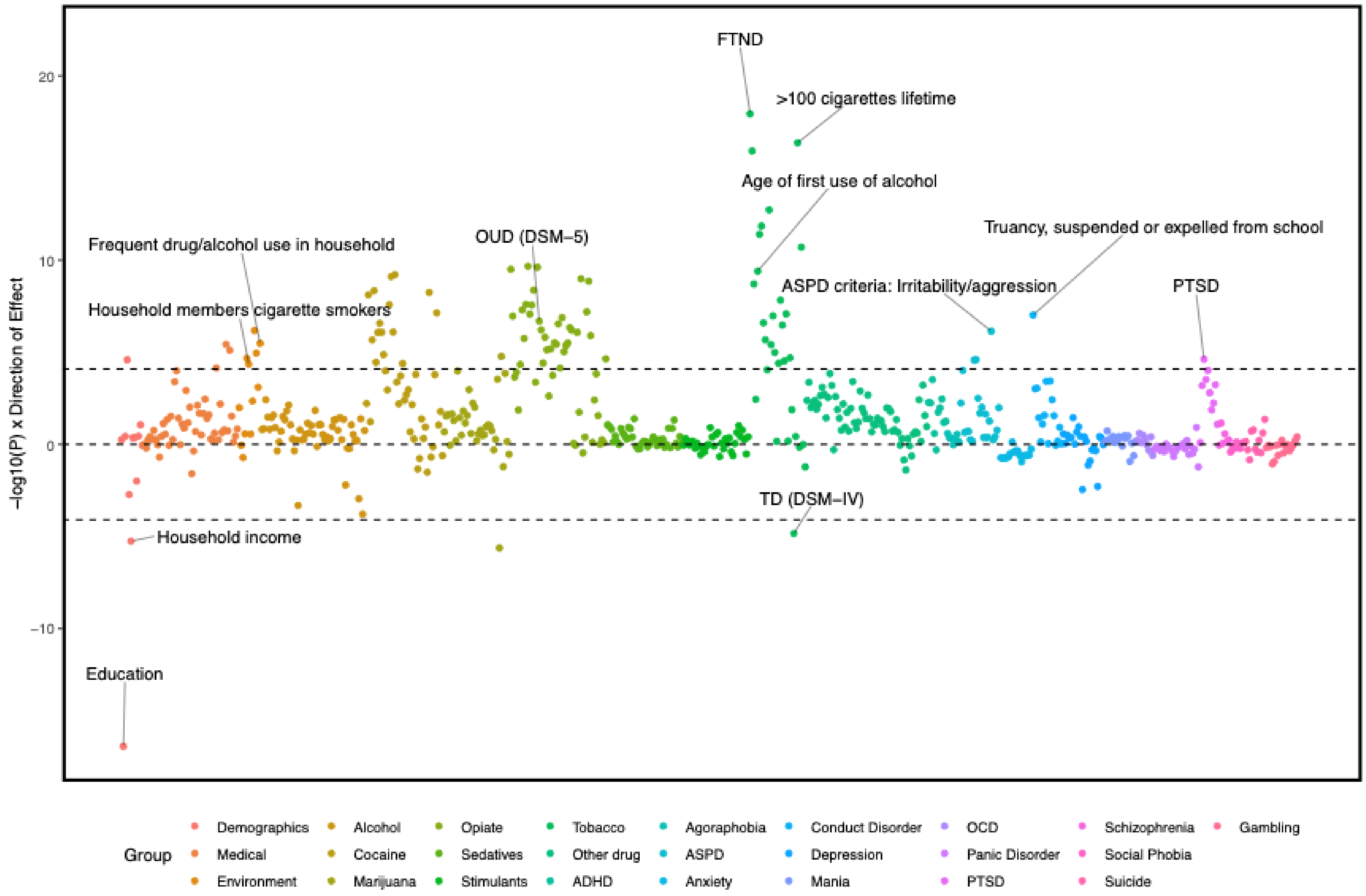

Supplementary Figure 6: $PGS_{CAD}$ covarying for CAD

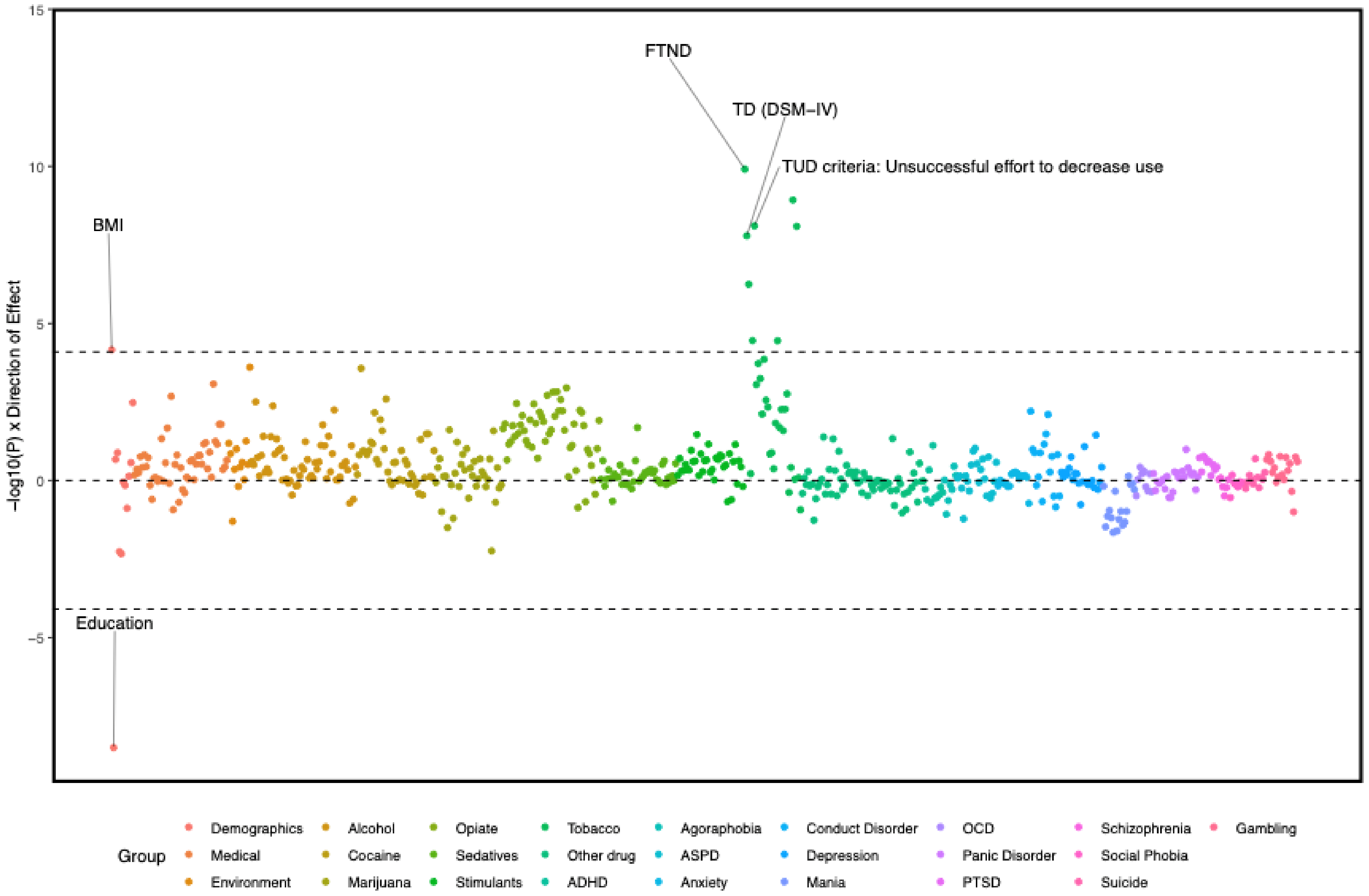

Supplementary Figure 7: $PGS_{T2D}$ covarying for T2D

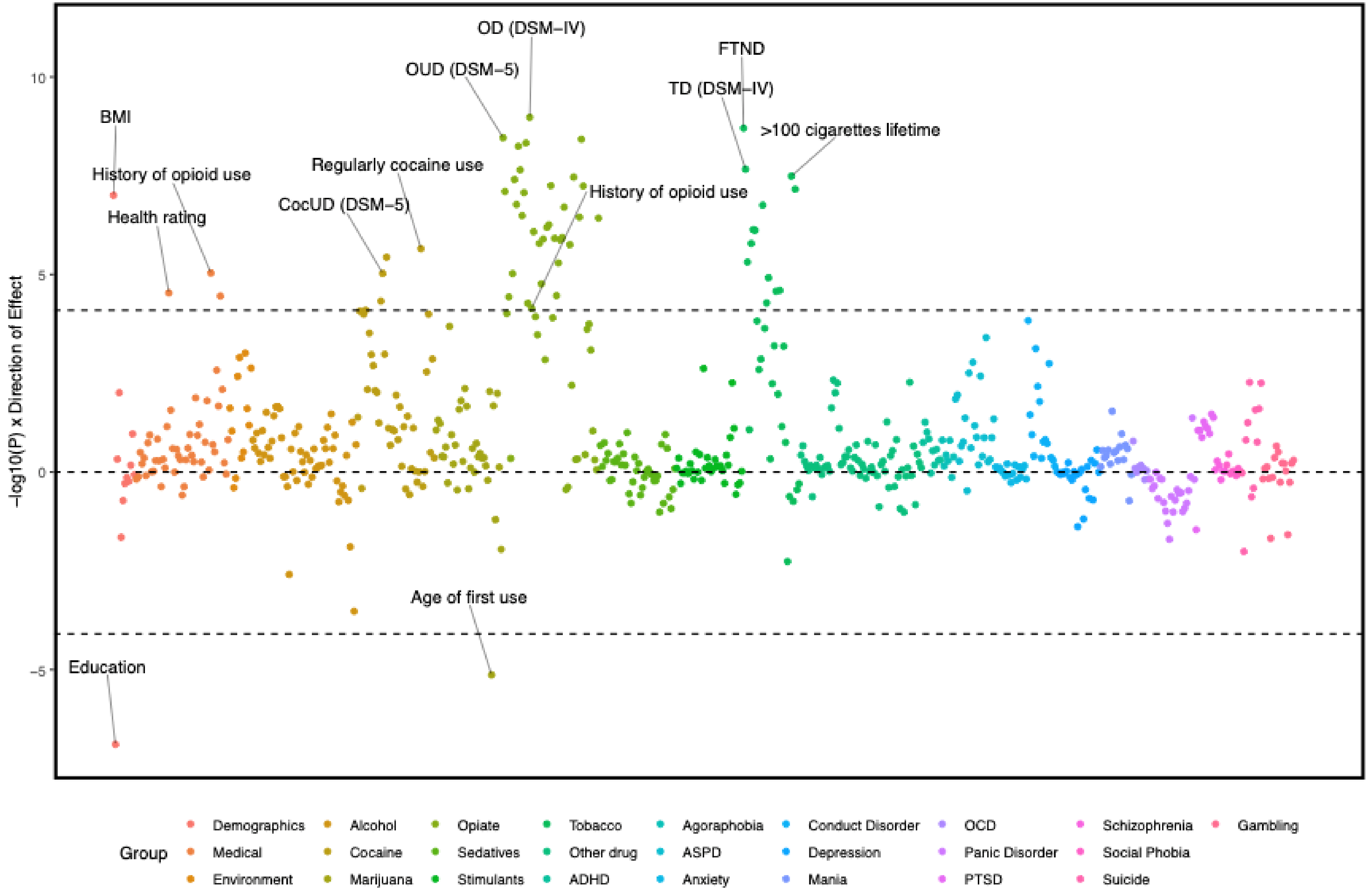
